# Characteristics of children readmitted with severe pneumonia in Kenyan hospitals

**DOI:** 10.1101/2024.02.21.24302816

**Authors:** Diana Marangu-Boore, Paul Mwaniki, Lynda Isaaka, Teresiah Njoroge, Livingstone Mumelo, Dennis Kimego, Achieng Adem, Elizabeth Jowi, Angeline Ithondeka, Conrad Wanyama, Ambrose Agweyu, the CIN Author Group

## Abstract

**Background:** Pneumonia is a leading cause of childhood morbidity and mortality. Hospital re-admission may signify missed opportunities for care or undiagnosed comorbidities.

**Methods:** We conducted a retrospective cohort study including children aged ≥2 months to 14 years hospitalised with severe pneumonia between 2013 and 2021 in a network of 22 primary referral hospitals in Kenya. Severe pneumonia was defined using the World Health Organization criteria, and re-admission was based on clinical documentation from individual patient case notes. We estimated the prevalence of re-admission, described clinical management practices, and modelled risk factors for re-admission and inpatient mortality.

**Results:** Among 20,603 children diagnosed with severe pneumonia, 2,274 (11.0%, 95% confidence interval (CI) 10.62 to 11.47) were readmitted. Re-admission was independently associated with age (12-59 months vs 2-11 months: adjusted odds ratio (aOR) 1.70, 95% confidence interval (CI) 1.55 to 1.88; >5 years vs 2-11 months: aOR 1.86, 95% CI 1.55 to 2.23), malnutrition (weight for age z-score (WAZ) < -3SD vs WAZ > -2SD: aOR 2.03, 95%1.83 to 2.28); WAZ -2 to -3 SD vs WAZ> -2SD: aOR 1.37, 95% CI 1.20 to 1.56) and presence of a concurrent neurological disorder (aOR 4.04, 95% CI 1.57 to 10.42) . Chest radiography was ordered more frequently among those readmitted (540/2,274 vs 3,102/18,329, p<0.001). Readmitted patients were more likely to receive second-line antibiotics (808/2,256 vs 5,538/18,173 p<0.001), TB medication (69/2,256 vs 298/18,173 p<0.001), salbutamol (530/2,256 vs 3,707/18,173 p=0.003), and prednisolone (157/2,256 vs 764/18,173 p<0.001). Inpatient mortality was 2,354/18,329 (12.8%) among children admitted with a first episode of severe pneumonia and 269/2,274 (11.8%) among those who were readmitted (adjusted hazard ratio (aHR) 0.94, 95% CI 0.82-1.07). Age (12-59 months vs 2-11 months: aHR 0.62, 95% 0.57 to 0.67), female sex (aHR 1.23, 95% 1.14 to 1.33), malnutrition (WAZ <-3SD vs WAZ> -2SD: aHR 1.90 95% CI 1.74 to 2.08); WAZ -2 to -3 SD vs WAZ> -2SD: aHR 1.48, 95% CI 1.32 to 1.65), incomplete vaccination (aHR 1.43, 95% CI 1.16 to 1.75), and anaemia (aHR 2.16, 95% CI 1.90 to 2.45) were independently associated with mortality.

**Conclusions:** Children readmitted with severe pneumonia account for a substantial proportion of pneumonia hospitalisations and deaths. Further research is required to develop evidence-based approaches to screening, case management, and follow-up of children with severe pneumonia, prioritising those with underlying risk factors for readmission and mortality.

## Background

In many low-and-middle-income-countries (LMICs), children and adolescents disproportionately face the double burden of acute respiratory infectious diseases and chronic comorbidities, resulting in long-term impairment of lung function (1, 2), reduced quality of life (3), and mortality (4). Presence of chronic comorbidities and prior severe pneumonia episodes increase the risk of pneumonia re-admission and is associated with substantial costs (5, 6). Almost 10% of children with community acquired pneumonia have recurrent pneumonia (7, 8). Recurrent pneumonia is defined as at least two episodes of pneumonia in one year or three episodes ever, with interval radiographic clearing of opacities; whereas persistent pneumonia is characterized by persistence of symptoms and radiographic abnormalities for more than one month (9). Recurrent and persistent pneumonia have varied aetiologies that are known antecedents to bronchiectasis or childhood interstitial lung disease (8, 10, 11), including rare genetic conditions that are increasingly being reported in sub Saharan Africa (12–14). Obstructive airway disease in children hospitalised with severe pneumonia may be underdiagnosed, and other differential diagnoses may not be considered (15, 16). While predictors of repeated acute hospitalisation for paediatric asthma which tends to co-exist with pneumonia are known and relatively modifiable, those for severe pneumonia readmission in sub-Saharan Africa are lacking (17).

The World Health Organization (WHO) guidelines for pneumonia used in many LMICs comprise a syndromic approach to disease classification and management to enable health workers at the first level referral hospitals to provide basic care for young children (18). However, the validity of the syndromic approach to pneumonia classification and management has yet to be tested among children presenting for severe illness after a previous episode of hospitalisation. Robust data on inpatient management of children with repeated hospitalisation for severe pneumonia are needed to guide appropriate policy recommendations on preventive, diagnostic, treatment and follow-up measures to optimize care and outcomes.

The aims of this study were to: i) establish the proportion of children aged 2 months to 14 years who were re-admitted with severe pneumonia in an established network of Kenyan public hospitals; ii) determine the factors associated with severe pneumonia re-admissions in this population; iii) describe the investigations and treatment provided to children re-admitted with severe pneumonia; and iv) explore the risk factors associated with mortality among these children.

## Methods

### Reporting

The reporting of this observational study follows the Strengthening of Reporting of Observational studies in Epidemiology (STROBE) statement (19).

### Ethics, consent and permissions

The Kenya Medical Research Institute (KEMRI) Scientific and Ethics Review Unit approved the collection of the deidentified data analysed in this study and waived the requirement for individual informed consent from patients whose data were used for this research. The Clinical Information Network (CIN) is run in collaboration with the Kenya Paediatric Association, Ministry of Health, and participating hospitals with the aim to improving routine clinical documentation for quality improvement, and for observational and interventional research (20).

### Study design and setting

We undertook a retrospective cohort study involving patients admitted to CIN hospitals. The CIN was initiated in October 2013 and comprises 22 purposefully selected county hospitals distributed across the densely populated central and western regions of Kenya (21, 22). Patients are admitted by a junior clinician under the supervision of one or more hospital paediatricians. CIN implements the use of structured clinical forms for all admitted children, capturing biodemographic data, caregiver-reported medical history, including whether the patient was readmitted and vaccination status according to the national routine schedule, clinical signs, diagnoses, diagnostic tests ordered, treatment plan, and inpatient outcome. Decisions on clinical and nursing management are made at the discretion of hospital staff with access to basic laboratory and radiological services. Case management is based on the Kenya national paediatric guidelines (23) adapted from WHO guidelines (18) as a reference. Patients are reviewed at least once daily for clinical progress. At hospital discharge, caregivers receive instructions on correct administration of continuing treatments and a follow up outpatient clinic visit is scheduled typically within two weeks.

### Study participants, data sources and variables

The study population comprised all children admitted to paediatric wards in CIN hospitals. Children hospitalised with a documented diagnosis of pneumonia were included. Children aged less than 2 months or more than 14 years, and those without severe pneumonia defined using the World Health Organization criteria between 2013 and 2021 were excluded. Trained data clerks extract deidentified individual patient data from routine hospital records into a REDCap upon discharge (22, 24). For this analysis, data retrieved comprised: the patient’s unique identifier, age, sex, hospital unique identifier, prior pneumonia admission (we could not compute the time from the first admission given the nature of the data), known comorbidities or risk factors such as malnutrition, anaemia, chronic neurological disorder, asthma, cardiac disease, renal disease, HIV, TB and sickle cell disease. We also extracted clinical data comprising management instituted during hospitalisation such as routine laboratory investigations e.g. full blood count; plain chest radiography; supportive treatment e.g. oxygen supplementation; antimicrobials treatment (first line, second line, third line antibiotics, coverage for atypical pathogens, antitubercular and antifungal medications); inhaled or systemic corticosteroids; and outcomes including discharge, referral or death. It should be noted that the structured forms used in CIN to document diagnostics and treatments only record affirmative entries (i.e., “Yes,” “Ordered,” or “Prescribed”). As a result, we inferred that entries which were not recorded on the forms were negative.

### Statistical analysis

Data were exported to STATA (StataCorp. 2019. *Stata Statistical Software: Release 16*.1 College Station, TX: StataCorp LLC.) for statistical analysis. Descriptive statistics were summarized as frequencies with percentages for categorical data, means with standard deviations for continuous data that were normal and medians with interquartile range for continuous data that were skewed. We estimated the proportion of severe pneumonia re-admissions among children aged 2 months to 14 years hospitalised within the CIN.

A multivariable mixed-effects logistic model was fitted to determine factors associated with re-admission among patients with severe pneumonia in this clinical cohort and we reported adjusted risk ratios. CIN data are hierarchical because admission episodes are nested within 22 hospitals, thus we used a random intercept logistic regression model to account for clustering (25). In the formula that follows, Y_ij_ denotes pneumonia readmission for the i^th^ child admitted in the j^th^ hospital (Y*_ij_* = 1 denotes pneumonia readmission, while Y*_ij_* = 0 denotes no pneumonia readmission); X*_1ij_*, through X*_kij_* denote the k explanatory variables measured on the i^th^ child admitted in the j^th^ hospital (e.g., child’s age); and *α*_0*j*_ denotes the unobserved cluster effect. Our model had no hospital level variables. It incorporates cluster-specific random effects to account for the within-cluster correlation of patient outcomes.

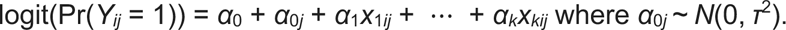

The assumption is made that the random effects are independent of the model covariates (X). Explanatory variables included age, sex, weight-for-age z-score (WAZ), vaccination status, previous wheeze and comorbidities including anaemia, HIV, TB, and neurological disorders. The Stata command “or:xtmelogit” was used to fit the random intercept model.

To determine factors associated with mortality among children with severe pneumonia readmissions, we fitted a multivariable mixed-effects Cox proportional hazard model (shared gamma frailty model) and reported adjusted hazard ratios with corresponding 95% confidence intervals. In the formula that follows, α_j_ denotes the random effect associated with the j-th hospital. The shared frailty term has a multiplicative effect on the baseline hazard function: h_i_ (t) = h_0_(t) exp(α_j_) exp(**X**_j_ β) (26). Explanatory variables included age, sex, weight-for-age z-score (WAZ), vaccination status, previous wheeze and comorbidities including anaemia, HIV, TB, and neurological disorders. We generated Cox proportional hazard regression plots for survival by type of pneumonia admission (first episode pneumonia and pneumonia re-hospitalisation); by age group (under 12 months, 12-59 months and over 5’s), gender, WAZ category (<-3 SD, -2 to -3 SD, >-2SD), vaccination and anaemia status. The Stata command “stcox” was used to fit the shared frailty Cox proportional hazard model. Censoring occurred at the time of death, discharge or referral, whichever came first.

For all models, explanatory variables were selected *a priori* based on existing literature, clinical relevance with regard to effect on outcome, ease of identification for risk stratification, and biological plausibility. P-values were rounded to three decimal places and were deemed statistically significant if they were <0.05 (2-sided). To handle missing data, we performed multiple imputation by chained equations generating 10 imputed datasets under the assumption that data were missing at random (27).

## Results

Out of 49,872 children admitted with pneumonia during the study period, 29,269 (59%) were excluded from the analysis. Exclusions were 27,977 children who did not have severe pneumonia, and 1,292 children aged < 2 months or > 14 years. Figure 1 shows the study population inclusion process. The main variables with missing values were gender, which was missing for 127 children (0.62%), and vaccination status which was missing for 13,468 children (65%).

**Figure 1:**
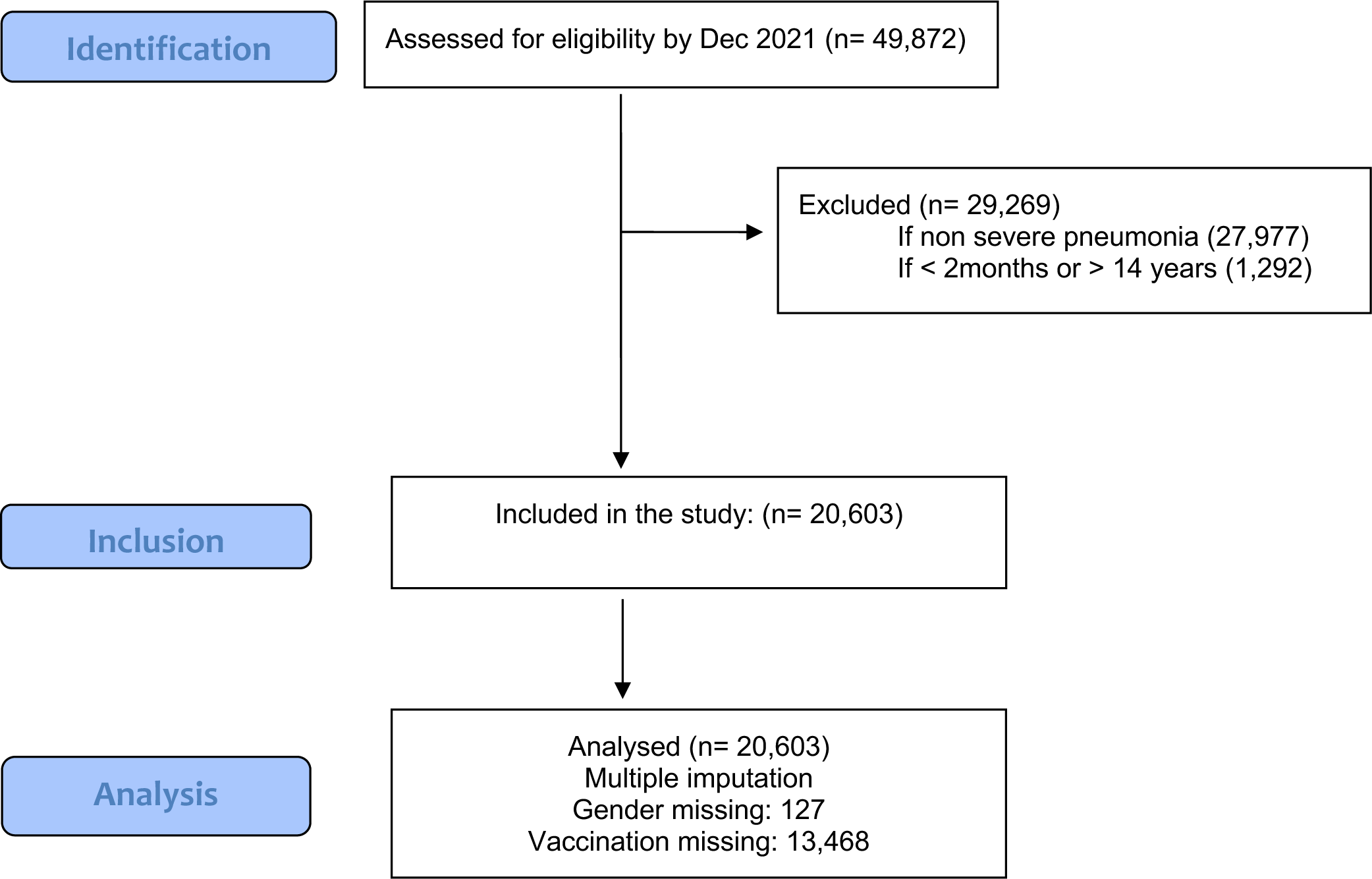
STROBE Flow Chart. Flow diagram of eligible study participants.

### Proportion of children re-admitted with severe pneumonia

Of the population included in the analysis comprising 20,603 children aged 2 months to 14 years hospitalised with severe pneumonia over the study period, 2,274 were readmitted, giving an estimated prevalence of 11.0% (95% 10.62-11.47) for readmissions among children with severe pneumonia.

### Characteristics of study participants

The median age of the study population was 12 months (IQR 6.0-24.0), 9,907/20,603 (48%) of children were under 12 months of age, 9,287/20,603 (45%) of the children were aged 12-59 months and 1,409/20,603 (6.8%) were aged 5-14 years. The majority of children were reported to be up to date with their vaccination status 6,950/7,135 (97%). However, most were severely or moderately underweight 17,018/20,603 (83%). Compared to children admitted for the first time with severe pneumonia, children re-admitted with severe pneumonia were slightly older (median age 14.4 months vs 12 months), underweight (median WAZ score -1.7 vs -1.1), were diagnosed with TB (1.8% vs 0.7%), and neurological disorders (0.4% vs 0.12%) more frequently (Table 1).

**Table 1:** Admission characteristics of study participants.

| Indicator | Levels | Total | Severe<br>Pneumonia<br>1<sup>st</sup> Admission | Severe<br>Pneumonia<br>Readmission | p-value |
| --- | --- | --- | --- | --- | --- |
|  |  | 20,603 | 18,329 | 2,274 |  |
| Age months Median<br>(IQR) |  | 12.0 (6.0-24.0) | 12.0 (6.0-24.0) | 14.4 (9.6-28.8) | <0.001 |
|  | Missing | 0 | 0 | 0 |  |
| (%) | <12mo | 9,907 (48.1) | 9,078 (49.5) | 829 (36.5) | <0.001 |
|  | 12-59mo | 9,287 (45.1) | 8,030 (43.8) | 1,257 (55.3) |  |
|  | Over 5yrs | 1,409 (6.8) | 1,221 (6.7) | 188 (8.3) |  |
|  | Missing | 0 | 0 | 0 |  |
| Gender (%) | Male | 11,334 (55.4) | 10,049 (55.2) | 1,285 (56.8) | 0.13 |
|  | Missing | 127 (0.62) | 114 (0.63) | 13 (0.57) |  |
| WAZ Median (IQR) |  | -1.1(-2.46, -0.46) | -1.1(-2.40, -0.03) | -1.7(-3.25, -0.51) | <0.001 |
|  | Missing | 812 | 718 | 94 |  |
| (%) | <-3SD | 14,225 (69.0) | 12,914 (70.5) | 1,311 (57.7) | <0.001 |
|  | -2 to -3SD | 2,793 (13.6) | 2,443 (13.3) | 350 (15.4) |  |
|  | >-2SD | 3,584 (17.4) | 2,972 (16.2) | 613 (27.0) |  |
|  | Missing | 0 | 0 | 0 |  |
| Vaccination | Up to date | 6,950 (97.4) | 6,145 (97.5) | 805 (96.9) | 0.3 |
|  | Missing | 13,468 (65.37) | 12,025 (65.61) | 1,443 (63.46) |  |
| Wheeze | Present | 3,240 (16.2) | 2,874 (16.1) | 805 (16.6) | 0.54 |
|  | Missing | 545 (2.65) | 475 (2.59) | 70 (3.08) |  |
| Anaemia | Present | 1,000 (4.9) | 909 (5.0) | 91 (4.0) | 0.045 |
|  | Missing | 0 | 0 | 0 |  |
| HIV | Positive or Exposed | 231 (1.1) | 197 (1.1) | 34 (1.5) | 0.073 |
|  | Missing | 1 | 1 | 0 |  |
| TB | Positive | 161 (0.8) | 120 (0.7) | 41 (1.8) | <0.001 |
|  | Missing | 0 | 0 | 0 |  |
| Neurological disorder | Present | 22 (0.1) | 12 (0.01) | 10 (0.4) | <0.001 |
|  | Missing | 0 | 0 | 0 |  |
| Saturations <90% |  | 7,717 (37.5) | 6,946 (37.9) | 771 (33.9) | <0.001 |

WAZ weight-for-age Z-score; Abnormal WAZ <-2SD
Neurological disorder: cerebral palsy, convulsive disorder or epilepsy

### Factors associated with severe pneumonia re-admissions

Age (12-59 months vs 2-11 months: adjusted odds ratio (aOR) 1.70, 95% 1.55 to 1.88), over 5 years vs 2-11 months: aOR 1.86, 95% 1.55 to 2.23), severe malnutrition (WAZ <-3SD vs WAZ> -2SD) (aOR 2.03, 95%1.83 to 2.28), moderate malnutrition (WAZ <-3SD vs WAZ> - 2SD) (aOR 1.37, 95%1.20 to 1.56), and neurological disorder (aOR 4.04, 95% 1.57 to 10.42) were independent risk factors for re-admission adjusting for gender, vaccination status, any wheeze, anaemia, HIV disease and TB (Table 2).

**Table 2:** Risk factors for readmission among study participants.

| Factor | Adjusted odds ratio<br>(95% confidence interval) | p-value |
| --- | --- | --- |
| Age (ref: <12 months) |  |  |
| 12-59 months | 1.70 (1.55-1.88) | <0.0001 |
| Over 5yrs | 1.86 (1.55-2.23) | <0.0001 |
| Gender (ref: female) | 1.04 (0.95-1.14) | 0.372 |
| WAZ (ref: >-2SD) |  |  |
| -2 to -3SD | 1.37 (1.20-1.56) | <0.0001 |
| <-3SD | 2.04 (1.83-2.28) | <0.0001 |
| Vaccination (ref: incomplete) | 0.74 (0.50-1.08) | 0.116 |
| Wheeze (ref: absent) | 1.49 (0.63-3.52) | 0.367 |
| Anaemia (ref: absent) | 0.96 (0.76-1.22) | 0.744 |
| HIV (ref: negative) | 1.15 (0.77-1.72) | 0.492 |
| TB (ref: absent) | 1.36 (0.91-2.02) | 0.129 |
| Neurological disorder (ref: absent) | 4.04 (1.57-10.42) | 0.004 |

Number of imputations=10; Number of observations=20,602; Number of groups=20;
Outcome = severe pneumonia readmission.
Imputed variables = gender 127; vaccination 13,468.
WAZ weight-for-age Z-score

### Investigations and treatment provided to study participants

Compared to children admitted for the first time with severe pneumonia, children re-admitted with severe pneumonia had chest radiography performed more frequently (24% vs 17%, p<0.001); and more prescriptions for second line antibiotics, specifically ceftriaxone (36% vs 30%, p<0.001), TB treatment (3% vs 1.6%, p<0.001), salbutamol (23% vs 20%, p=0.003) and prednisone (7% vs 4%, p<0.001) (Table 3).

**Table 3:** Diagnostics and treatments used among study participants.

|  |  | Total(N=20,603) | Severe Pneumonia<br>First Admission (N=18,329) | Severe Pneumonia<br>Readmission (N=2,274) | p-value |
| --- | --- | --- | --- | --- | --- |
| INVESTIGATIONS (%) |  |  |  |  |  |
| Pulse oximetry done | Yes | 13,245 (64.3) | 11,940 (65.1) | 1,305 (57.4) | <0.001 |
|  | No | 7,358 (35.7) | 6,389 (34.9) | 969 (42.7) |  |
| Chest radiography | Yes | 3,642 (17.7) | 3,102 (16.9) | 540 (23.8) | <0.001 |
|  | No | 16,961 (82.3) | 15,227 (83.1) | 1,734 (76.3) |  |
| TREATMENTS (%) |  |  |  |  |  |
| Oxygen ordered | Yes | 5,839 (28.3) | 5,216 (28.5) | 623(27.4) | 0.540 |
|  | No | 14,764 (71.7) | 13,113 (71.5) | 1,651 (72.6) |  |
| First line antibiotics |  |  |  |  |  |
| Crystalline penicillin | Yes | 16,376 (79.5) | 14,721 (80.3) | 1,655 (72.8) | <0.001 |
|  | No | 4,227 (20.5) | 3,608 (19.7) | 619 (27.2) |  |
| Gentamicin | Yes | 14,613 (70.9) | 13,233 (72.2) | 1,380 (60.7) | <0.001 |
|  | No | 5,990 (29.1) | 5,096 (27.8) | 894 (39.3) |  |
| Amoxicillin | Yes | 1,220 (5.9) | 1,104 (6.0) | 116 (5.1) | 0.20 |
|  | No | 19,383 (94.1) | 17,225 (94.0) | 2,158 (94.9) |  |
| Second line antibiotics |  |  |  |  |  |
| Ceftriaxone | Yes | 6,346 (30.8) | 5,538 (30.2) | 808 (35.5) | <0.001 |
|  | No | 14,257 (69.2) | 12,791 (69.8) | 1,466 (64.5) |  |
| Other antimicrobials |  |  |  |  |  |
| Metronidazole | Yes | 1,002 (4.9) | 894 (4.9) | 108 (4.7) | 0.92 |
|  | No | 19,601 (95.1) | 17,435 (95.1) | 2,166 (95.3) |  |
| Cotrimoxazole | Yes | 538 (2.6) | 480 (2.6%) | 58 (2.6) | 0.94 |
|  | No | 20,065 (97.4) | 17,849 (97.4) | 2,216 (97.4) |  |
| TB medication | Yes | 367 (1.8) | 298 (1.6%) | 69 (3.0) | <0.001 |
|  | No | 20,236 (98.2) | 17,875 (98.4) | 2,205 (97.0) |  |
| Other treatment |  |  |  |  |  |
| Salbutamol | Yes | 4,237 (20.6) | 3,707 (20.2) | 530 (23.3) | 0.003 |
|  | No | 16,366 (79.4) | 14,622 (79.8) | 1,744 (76.7) |  |
| Prednisolone | Yes | 921 (4.5) | 764 (4.2%) | 157 (6.9) | <0.001 |
|  | No | 19,682 (95.5) | 17,565 (95.8) | 2,177 (93.1) |  |

### Outcomes of study participants

The median length of hospital stay was 4 days IQR (2–7) for children hospitalised with a severe pneumonia for the first time; and 5 days IQR (3–8) for children readmitted with severe pneumonia. We noted that children readmitted with severe pneumonia were hospitalised for longer than seven days more frequently than those admitted for the first time with severe pneumonia (34% vs 26%, p<0.001). Among 18,329 children admitted with severe pneumonia for the first time, 2,354 (12.8% [12.37-13.33]) died during their hospital stay, while 269 (11.8% [10.56-13.22]) of 2,274 children readmitted with severe pneumonia died during their hospital stay (Table 4).

**Table 4:** Outcomes of study participants.

|  |  | Total(N=20,603) | Severe Pneumonia<br>First Admission (N=18,329) | Severe Pneumonia<br>Readmission (N=2,274) | p-value |
| --- | --- | --- | --- | --- | --- |
| Length of stay |  |  |  |  |  |
| Median [IQR] (Range) days |  | 4 [2-7] (1,401) | 4 [2-7] (1,339) | 5 [3-8] (1,401) |  |
| Missing |  | 1 | 1 | 0 |  |
| Less than 7 days |  | 15,159 (73.6) | 13,650 (74.5) | 1,509 (66.4) | <0.001 |
| 7 days or more |  | 5,444 (26.4) | 4,679 (25.5) | 765 (33.6) |  |
| Missing |  | 0 | 0 | 0 |  |
| Discharged |  | 17,261 (83.8) | 15,337 (83.7) | 1,924 (84.6) | 0.18 |
| Referred |  | 641 (3.1) | 567 (3.1) | 74 (3.3) |  |
| Died |  | 2,623 (12.7) | 2,354 (12.8) | 269 (11.8) |  |
| Missing |  | 78 (0.4) | 71 (0.4) | 7 (0.3) |  |

### Risk factors associated with mortality

Age <12 months (adjusted HR (aHR) 1.63, 95% 1.49 to 1.77), age >5 years (aHR 1.52, 95% 1.31 to 1.77), female gender (aHR 1.23, 95% 1.14 to 1.33), severe malnutrition (aHR 1.90 95% 1.74 to 2.08), moderate malnutrition (aHR 1.48, 95% 1.32 to 1.65), incomplete vaccination (aHR 1.43, 95% 1.16 to 1.75) and anaemia (aHR 2.16, 95% 1.90 to 2.45) were independently associated with mortality amongst children admitted with severe pneumonia when adjusted for gender, any wheeze, HIV, TB and a neurological disorder (Table 5 and Figure 2).

**Table 5:** Factors associated with mortality among children admitted with severe pneumonia in Kenyan hospitals within the Clinical Information Network (CIN)

| Factor | Adjusted hazard ratio (95% CI) | p-value |
| --- | --- | --- |
| Pneumonia re-admission (ref: first episode pneumonia) | 0.94 (0.82-1.07) | 0.338 |
| Age (ref: <12 months) |  |  |
| 12-59 months | 0.62 (0.57-0.67) | <0.0001 |
| Over 5yrs | 0.94 (0.81-1.08) | 0.379 |
| Gender (ref: male) | 1.23 (1.14-1.33) | <0.0001 |
| WAZ (ref: >-2SD) |  |  |
| -2 to -3SD | 1.48 (1.32-1.65) | <0.0001 |
| <-3SD | 1.90 (1.74-2.08) | <0.0001 |
| Vaccination not up to date (ref: complete) | 1.43 (1.16-1.75) | 0.001 |
| Wheeze present (ref: absent) | 0.21 (0.03-1.50) | 0.120 |
| Anaemia present (ref: absent) | 2.16 (1.90-2.45) | <0.0001 |
| HIV infected/exposed (ref: negative) | 1.10 (0.84-1.45) | 0.483 |
| TB positive (ref: absent) | 1.08 (0.74-1.58) | 0.674 |
| Neurological disorder (ref: absent) | 1.48 (0.55-3.96) | 0.219 |

WAZ weight-for-age Z-score

**Figure 2:**
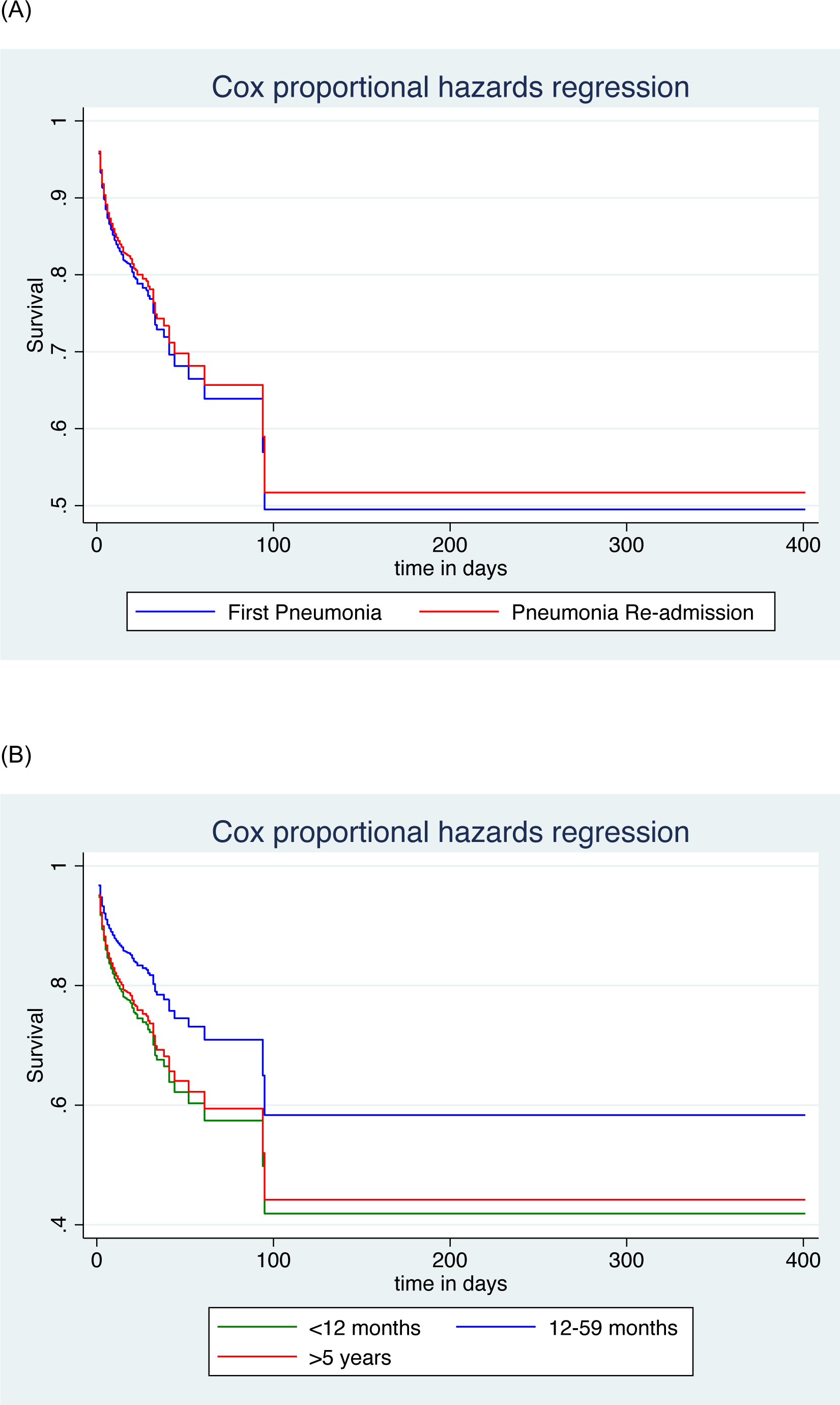
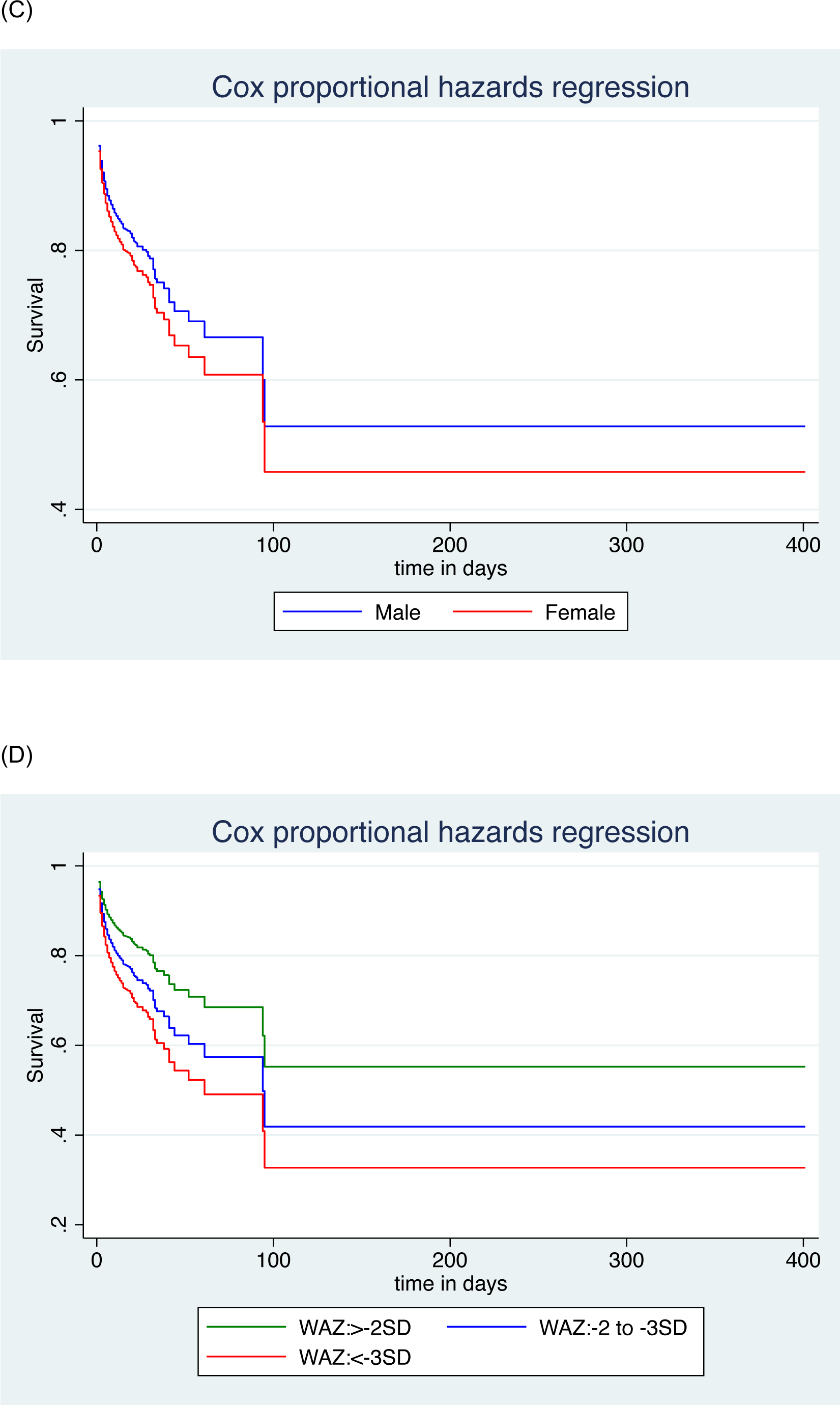
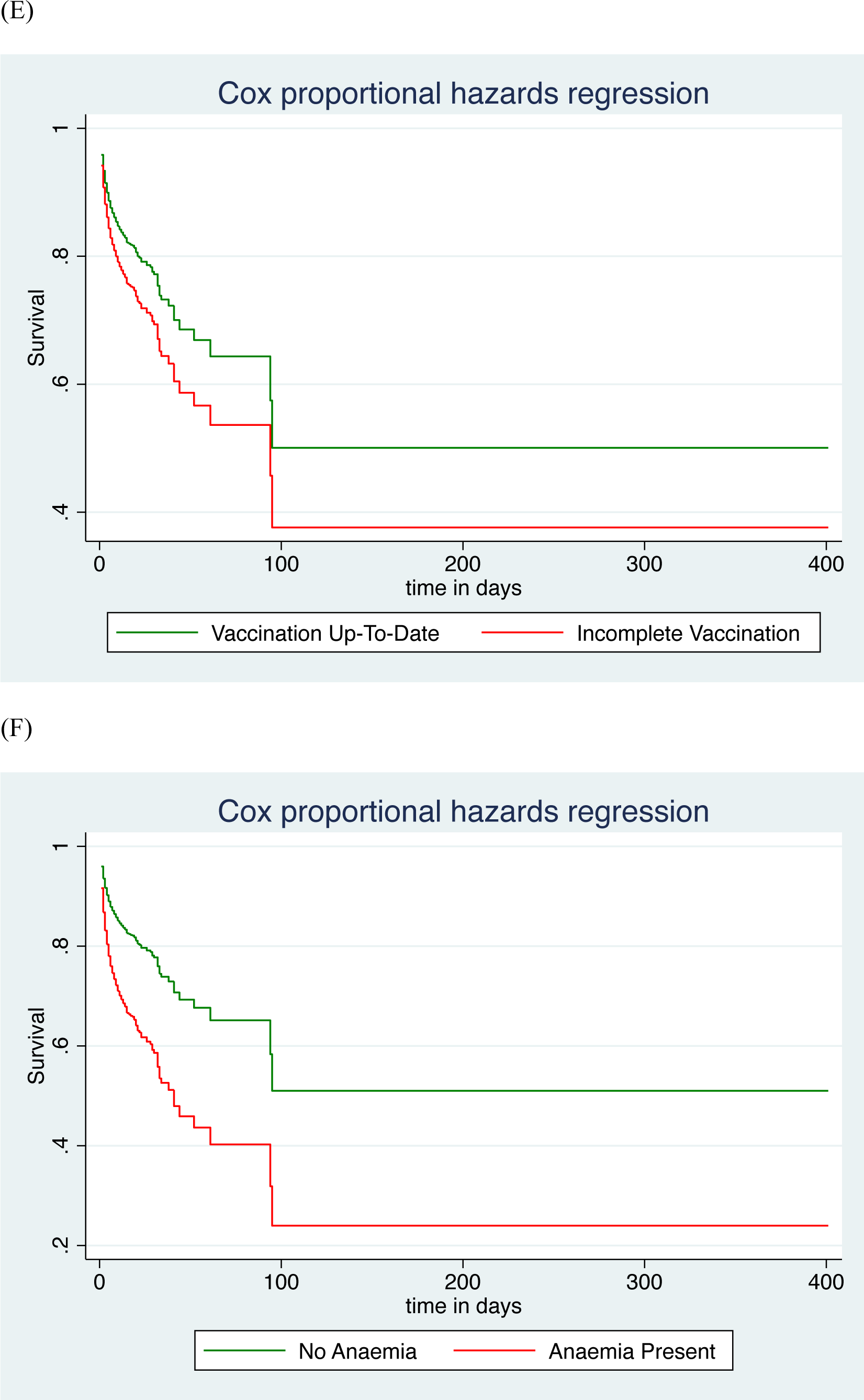
Cox proportional hazard plots for mortality among study participants.

## Discussion

Our analysis revealed a high proportion of children aged 2 months to 14 years with severe pneumonia in Kenyan secondary-level health facilities had previously been admitted for care. Risk factors for re-hospitalisation for severe pneumonia are age >12months, moderate/severe undernutrition and neurological disorder. The proportion of children getting plain chest radiography, second line antibiotics, TB medication, salbutamol and prednisone was higher among those re-admitted compared to those hospitalised for the first time. Mortality among children hospitalised with severe pneumonia is high, but not significantly different among those readmitted compared to first admissions. Children who are <12 months, over 5 years, female, with moderate/severe undernutrition or anaemia are at a higher risk of dying.

Publications on severe pneumonia readmission in children in LMICs are limited. Conversely, financial incentives and penalties imposed by hospital readmission reduction programs seem to drive such studies in high income countries (HICs), more so among adults (28). Re-admissions can be classified as early (≤30 days) or late (≥31 days) (29). Re-admissions that occur within 30-days often correspond to those that are potentially avoidable, most of which are direct or indirect complications of underlying comorbidities (30). These are the readmissions health facilities and insurance companies are keen on reducing, and which hospitals in HICs are subject to penalties if unacceptable thresholds are reached (31). Large longitudinal studies of various tertiary referral hospitals in the United States of America (USA), reported a 30-day lower respiratory infection specific re-admission rate and paediatric pneumonia specific re-admission rate of 2.7% (32) and 3.3% (5) respectively, almost four times lower than in our study. A more recent analysis conducted in the USA using the 2018 Nationwide Readmissions Database reported a 30-day severe pneumonia specific re-admission rate among children of 8.7%, while the re-admission rate in children with non-severe pneumonia was 4.7% (33). The lower estimates found in the USA may be attributed to the difference in income contexts, pneumonia definitions, and the fact that these were strictly 30-day outcomes. All the children in our analysis had severe pneumonia based on WHO guidelines. Furthermore, we could only ascertain that they had a prior hospital admission but we could neither determine the reason for the initial admission nor compute the time from the first admission given the nature of the data. It is imperative to obtain this time variable in future studies to report on comparable data with regard to early and late pneumonia readmissions.

Hospital re-admission may signal failure in the health system performance. However, its interpretation may be complicated by observed and unobserved differences in the clinical risk of patient populations and their survival rates. For example, health facilities with low childhood mortality may have a higher unobserved volume of sick children at risk of re-admission (34). In our study, mortality among children hospitalised with severe pneumonia was high, but not significantly different among those readmitted compared to first admissions. Indeed, hospital re-admissions are a more complex phenomenon than mortality, in that they arise from an “interplay among patient-, hospital-, community-, and environmental-level factors” (35). Comorbidities including neurological disorders and malnutrition, were risk factors for re-hospitalisation for pneumonia similar to our study (5, 32). In a cross-sectional study conducted at a Canadian tertiary hospital including almost 3,000 children, Owayed et al reported that most children hospitalised with recurrent pneumonia have a predisposing factor, the most common being oropharyngeal incoordination (36). This is a plausible explanation for the four-fold increased risk with severe pneumonia re-hospitalisation observed in children with a neurological disorder in our study. It calls for a high index of suspicion among clinicians from the first point of patient contact and consideration of medical or surgical interventions early. Immunosuppressive conditions have also been identified as a risk factor for pneumonia readmission in children (33). Immunosuppression can be due to moderate/severe malnutrition which was significantly associated with readmission in this study. However, in this analysis, HIV was not associated with severe pneumonia readmission; perhaps due to timely initiation of antiretroviral therapy within prevention of mother-to-child transmission services (37). Patient data on other conditions that present with recurrent pneumonia such as primary immunodeficiency, cystic fibrosis and primary ciliary dyskinesia that can lead to bronchiectasis (38) were not available. Unlike our study, age less than 1 year (5, 32) and male gender (32) have been identified as risk factors for re-hospitalisation for pneumonia. This could reflect differences in our study population. Perhaps, older children living longer to have a pneumonia readmission; or no actual particular gender risk for pneumonia rehospitalisation. Prior respiratory syncytial virus lower respiratory tract infection (39), longer index hospitalisations (5, 33), complicated pneumonia (5), and hospital case volume and teaching status (33) were other risk factors for re-hospitalisation for pneumonia identified in the literature but were not measured in this study. These are important variables that need to be examined in future studies.

Interestingly, Baseer et al estimated that 11.4% of children admitted with pneumonia during a one-year study in Egypt had recurrent pneumonia defined as ≥2 episodes of radiographically confirmed pneumonia in a single year or ≥3 episodes at any time with a radiographic clearing of densities between attack (40). In this analysis, almost 20% of children had a chest radiograph, with a significantly higher frequency of requests among children who were readmitted compared to those admitted with pneumonia for the first time. Data on interval assessment of chest radiographs was not available. It may be useful in future studies to document chest radiography findings and ascertain if children readmitted with severe pneumonia have recurrent pneumonia/persistent pneumonia. Timely interventions could potentially reverse, prevent or better manage chronic lung disease (8, 41).

Similar to our findings, other studies conducted in similar epidemiological and geographical contexts, specifically Kilifi at the Kenyan coast, Bangladesh and India, have found malnutrition (42–44) and incomplete vaccination (45) to be associated with mortality in children with severe pneumonia. We found severe pallor to be associated with mortality in children severe pneumonia (44, 46), consistent with previous findings among children with non-severe pneumonia (6). This association in children with pneumonia may be indicative of a reduced threshold for respiratory decompensation in this vulnerable population. Timely identification and management of anaemia is needed to prevent death in children hospitalised with severe pneumonia. Previous studies have described the association between higher mortality and infants with severe pneumonia (47), which we also found in our analysis. Interestingly, mortality in children over 5 years was similar to that observed among infants. Mortality among children aged 5-14 years has received little attention (48), with those in sub-Saharan Africa faring worse (49). Older children may have attendant complex comorbidities that put them at risk for death. Evidence-based treatment algorithms for this neglected population are needed. Moderate or severe malnutrition and neurological disorders in children with severe pneumonia should signal a need for post-admission follow up and potential re-admission. Optimizing nutrition may avert re-admission with severe pneumonia. Going forward, it will be important to conduct studies that objectively assess possible aspiration/reflux in children with neurological disorders readmitted with pneumonia in these settings perhaps using simple questionnaires at district-level health facilities (50) or/and more elaborate tests such as videofluoroscopic swallowing studies to assess swallowing (51) and scintigraphy, pH-metry and other studies to quantify gastro-oesophageal reflux in referral centres (52). Establishing the multi-disciplinary capacity necessary to improve care for children with neurologic disorders, specifically training physiotherapists and speech therapists is an urgent requirement.

In this study, referral of patients with severe pneumonia who had been readmitted was quite low at 3%, similar to those with a first hospitalisation with pneumonia. It is difficult to ascertain whether clinicians are reluctant to refer patients; or whether patients would honour the referral instructions. Exploring referral systems in future studies may prompt changes in patient pathways for children with severe pneumonia who have been re-hospitalised. This may inform the health facilities on the areas that could be improved, which could include increasing the number of specialists and sub-specialists in the counties through models such as the African Paediatric Fellowship Program (53). Remote consultation (54), among other solutions that adopt a contextually appropriate reasoned diagnostic approach for children with recurrent/persistent pneumonia may be considered (8).

To the best of our knowledge, this is the first published study to report on severe pneumonia rehospitalisation in children in sub-Saharan Africa. The large sample size across multiple settings provides a contemporary representation of paediatric severe pneumonia re-admission in a high burden context, offering a foundation for research and interventions averting or mitigating the clinical sequelae of pneumonia. The study is limited by its retrospective nature and use of routinely collected data, which may result in missing data on variables of interest, such as the interval between first admission and subsequent readmissions, challenges linking records for patients with multiple episodes of readmission, specialised modalities of testing employed (e.g., videofluoroscopy, echocardiography), and relevant complications (e.g., pleural effusions, abscess, air leaks).

## Conclusion

The proportion of children re-hospitalised with severe pneumonia and their in-hospital mortality is high. Age, nutritional status and neurological disorder are risk factors of severe pneumonia recurrence; additionally, gender, incomplete vaccination and anaemia increase risk of mortality. Optimal care for severe pneumonia will require carefully designed studies of clinical algorithms for the screening, case management, and follow up of children, prioritising those with underlying risk factors.

## List of abbreviations

aHR: adjusted Hazard Ratio
aOR: adjusted Odds Ratio
CIN: Clinical Information Network
KEMRI: Kenya Medical Research Institute
LMIC: Low-and-middle-income country
TB: tuberculosis
WAZ: Weight-for-Age Z-score
WHO: World Health Organization

## Declarations

### Ethics approval and consent to participate

The Kenya Medical Research Institute (KEMRI) Scientific and Ethics Review Unit approved the collection of the deidentified data analysed in this study. The CIN is run in collaboration with the Ministry of Health and participating hospitals with the aim to improve the quality of routine paediatric hospital data for use in audits, observational and interventional research. Individual consent for access to deidentified patient data was not required.

### Consent for publication

Not applicable.

### Availability of data and materials

The data utilized in this work was made available to the research team by the participating hospitals and the Ministry of Health, and thus we are not the primary data owners; our use for these routine hospital data is approved as part of a specific ethical review process. Further access to the data can be sought through a request to KEMRI Wellcome Trust Research Programmes’s Data Governance Committee through.

### Authors’ contributions

DM-B and AA conceived the study and DM-B performed the analysis with support from AA. DM drafted the initial manuscript with support from AA, PM, LI, TN, LM, DK, AA, EJ, AI, and CW. CIN authors contributed to the design of data collection tools, conduct of the work, collection of the data, and data quality assurance that form the basis of this manuscript. All authors read and approved the final version of the manuscript.

## Acknowledgements

The authors would like to thank the Ministry of Health who gave permission for this work to be developed and have supported the implementation of the Clinical Information Network (CIN) together with the County Health Executives, Hospital Management Teams, the Kenya Paediatric Association, the Kenya Ministry of Health, and the University of Nairobi for promoting the aims of the CIN. Specifically, teams based in: Busia County Hospital (Emma Sarah Namulala, Yuvane Maiyo); Bungoma County Hospital (Dickens Lubanga, Felistus Makokha); Homabay County Hospital (Meshack Liru, Edith Ogada); Vihiga County Hospital (Vitalis Juma); JOOTRH (Josephine Ojigo, Maureen Muchela); Kakamega CGTRH (Boniface Nyumbile, Roselyn Malangachi, Ijusa Midecha); Kitale County Hospital (Rachel Inginia, Paul J W Njanwe, Mwende Mutunga); Kiambu County Hospital (Grace Ochieng, Lydia Thuranira and Catherine Murianki); Mama Lucy Kibaki Hospital (Cecilia Mutiso, Celia Muturi, Elizabeth Atieno Jowi); Machakos County Hospital (Charles Nzioki, Penina Musyoka); Kisumu County Hospital (Achieng Adem, Sharon Ocharo); Embu County Hospital (Esther Mukami Njiru, Mwangi Ngina, Patrick Lydia); Naivasha County Hospitals (Julie Barasa); Thika Level 5 Hospital (Bernadette Lusweti, Maureen Njoroge, Sylvia Mwathi, Patrick Mburugu); Nakuru County Hospital (Angeline Ithondeka, Linda Ombito, Alice Nkirote Nyaribari, Elizabeth Kibaru); Kerugoya County Hospital (Peninah Muthoni Mwangi); Mbagathi County Hospital (Christine Manyasi, David Kimutai, Rukia Aden, Zanuba Mohammed); Nyeri County Hospital (Wagura Mwangi, Agnes Mithamo); Pumwani Maternity Hospital (Catherine Mutinda, Beth Maina, Rashid Musa, Caren Emadau, Maureen Muirithi, Esther Muthoni Ogolla); Migori County Hospital (Joyce Akuka); Malindi Sub-County Hospital (Yusuf Rasheed); Kisii County Hospital (Mourine Ikol); and Karatina Sub-County Hospital (Stella Mbuga).

The authors also acknowledge the other members of the CIN leadership: Jalemba Aluvaala, Anthony Etyang, Samuel Akech, Muthoni Ogola, Jacquie Oliwa, Michuki Maina, David Gathara, Fredrick Were, Grace Irimu, and Mike English, as well as technical and administrative assistance provided by Metrine Saisi, Elizabeth Muthini, James Wafula, Cynthia Khazenzi, Joyce Kigo, George Mbevi, Henry Gathuri, Wycliffe Nyachiro, and Frankline Oketch. This work is published with the permission of the Director of KEMRI CGMRC.

